# Investigating the spatial accessibility and coverage of the pediatric COVID-19 vaccine: an ecologic study of regional health data

**DOI:** 10.1101/2023.12.27.23300589

**Authors:** Amin Bemanian, Jonathan F Mosser

## Abstract

**Background:** The COVID-19 pandemic presented healthcare workers and public health agency a unique challenge of having to rapidly deliver a novel set of vaccines during a public health crisis. For pediatric patients, there was an additional layer of complexity given the delayed timeline to deliver these vaccines and the differences in dosing and available products depending on the age of those receiving the vaccine. This paper investigates the spatial accessibility and uptake of the COVID-19 vaccine in King County, WA, USA.

**Methods:** Public data for COVID-19 vaccine sites to calculate spatial accessibility using an enhanced two step floating catchment area (E2SFCA) technique. Spatial regression analyses were done looking at the relationship between spatial accessibility and ZIP code level vaccination rates. Relationships with other socioeconomic and demographic variables were calculated as well.

**Findings:** Higher rates of vaccine accessibility and vaccine coverage were found in adolescent (12 to 17-year-old) individuals relative to school age (5 to 11-year-old) individuals. Vaccine accessibility was positively associated with coverage in both age groups in the univariable analysis. This relationship was affected by neighborhood educational attainment.

**Interpretation:** This paper successfully demonstrates how spatial accessibility measures such as E2SFCA can be used to assess the availability of the COVID-19 vaccine in a region such as a metropolitan area or county. It also provides insight into some of the ecological factors that affect COVID-19 vaccination rates. Implementation of these technique could help public health authorities and healthcare organization plan future vaccination efforts.

## Introduction

Spatial accessibility to healthcare services is an important determinant of health, as it can affect a patient’s ability to receive preventative services, medications, and acute or critical care.

Within the field of pediatric medicine, there have been multiple studies showing how providers and other healthcare services are not evenly distributed across populations, with disparities across socioeconomic status, racial composition, and urban/rural areas.^1,3,4^ These differences in spatial accessibility have been shown to affect health outcomes as well. One study of routine immunizations among children with Medicaid insurance in Washington DC found that higher spatial accessibility to vaccination providers was associated with higher odds of routine vaccination completion.^5^ Spatial availability has been shown to affect the distribution of childhood vaccine doses as well. Mapping of human papillomavirus and tetanus/diphtheria/pertussis vaccine doses in Georgia was found to have spatial clustering at the county level, and the authors found that public transit and the number of health department clinics were positively associated with increased vaccine doses.^6^ These studies demonstrate the importance of spatial accessibility in pediatric health.

Ensuring accessibility of care during a pandemic, however, is particularly difficult. The COVID-19 pandemic caused significant strain to the United States medical system and changes to patients’ health seeking behaviors. Pediatric emergency department visits significantly decreased in 2020 relative to pre-pandemic levels and have slowly risen back over the course of 2021 and 2022.^7^ Additional work has shown that emergency department visits and hospitalizations have disproportionately decreased among children living in census tracts with lower socioeconomic scores as measured by the Child Opportunity Index.^8^ This decreased utilization has affected preventative care services as well, with routine childhood vaccinations across all age groups decreasing during the first year of the pandemic, especially dropping off during the initial months.^9^ Furthermore, the COVID-19 pandemic has presented a unique challenge of deploying novel vaccines to curb disease spread, while much of society continued to try to isolate. For these reasons, investigation into the accessibility of pediatric COVID-19 vaccine deployment is relevant to understanding how healthcare resource access has been distributed across population areas and where the greatest needs are for vaccine providers and resources.

This paper seeks to characterize the spatial accessibility of pediatric COVID-19 vaccination sites in King County, Washington (WA) in the United States. Furthermore, it identifies associations neighborhood-level determinants of health and vaccine accessibility to assess if there are specific communities that have limited access to vaccine providers. Finally, this study investigates the relationship between vaccine accessibility and coverage, specifically to test the hypothesis that increased accessibility is associated with higher rates of vaccine uptake within communities.

## Methods

The 2019 5-year American Community Survey (ACS) was used for demographic and socioeconomic data via the tidycensus R package.^10,11^ All variables were obtained at the ZIP Code Tabulation Area (ZCTA) level. Variables obtained from the ACS included age stratified population counts, the percentage of households living below the poverty line, the percentage of adults aged ≥25 years who graduated with a bachelor’s degree or higher, and the percentage of residents belonging to specific racial/ethnic groups. COVID-19 vaccine coverage data was obtained from Public Health - Seattle and King County (PHSKC) COVID-19 Dashboard reflecting the latest Washington Immunization Information System data up to 7/5/2022.^12^ Coverage was defined as the percentage of children who had completed a primary COVID-19 vaccination series (two doses of either the Pfizer-BioNTech BNT162b2 or Moderna mRNA-1273 mRNA vaccines). Vaccine coverage data was stratified by age into two groups: 5 to 11-year-olds (school age children) and 12 to 17-year-olds (adolescents). Data on younger children was not available at the time of this analysis.

Vaccine accessibility was estimated using the enhanced two-step floating catchment area (E2SFCA) technique.^13^ Vaccine providers within King County were identified using the public Washington Vaccine Locator website and were stratified based on which vaccine series they provided: the adolescent/adult series (n = 434) and/or the childhood series (n = 152). Any site that was listed as a “mobile center” was excluded from the analysis, due to a lack of information on whether the address corresponded to where the center distributed the vaccine or where the vaccines were stored between drives.

The steps of the E2SFCA technique are briefly described here. First, for every ZCTA, 200 spatial points within the ZCTA’s boundaries were randomly sampled, weighted based on a population density raster from WorldPop.^14^ This sampling process aimed to reduce bias from picking a single point (e.g. geometric centroid) to represent each ZCTA, which could particularly affect large, sparsely populated ZCTAs. Next, travel time distances from each sampled point were calculated to each vaccination site using the R5R package and OpenStreetMap road network files.^15^ Using these travel time distances, we then calculated the ZCTA’s accessibility score for each of the sampled points. Two sets of scores based on private automobile and public transportation times were calculated and then weighted by the percentage of households who did not own any private automobiles (from the American Community Survey) to calculate a combined score.^10,11^ From this sample space, the median accessibility score for each ZCTA was selected. The derivation of the time weighting function for the E2SFCA is provided in supplement S1. One ZCTA encompassing Vashon Island, an island municipality within King County, was excluded from this analysis due to requiring a ferry to reach the mainland. As a result, it was a significant outlier for vaccine accessibility scores.

Another ZCTA in the eastern part of the county was excluded as it crosses over into the neighboring Kittitas County. The relationship between accessibility and coverage was then assessed at the ZCTA level using spatial error regression with the spatialreg R package.^16,17^ Ordinary linear regression was unable to be used given spatial autocorrelation in the residuals of most of the models. Analysis was stratified by the two age groups. First, we assessed if vaccine accessibility was associated with household poverty, racial composition, and adult educational attainment at the ZCTA level. We then investigated the relationship between accessibility and coverage with several models: a series of univariable regressions with accessibility and the socioeconomic/demographic variables, a multivariable regression with the socioeconomic/demographic variables added as covariates with no interaction terms, and a multivariate regression which included significant interactions with vaccine accessibility. Finally, we stratified a univariable regression analysis between accessibility and coverage based on ZCTA educational attainment. We selected a threshold of 50% for adults with bachelor’s degrees (sample median: 52%) for the bifurcation. We conducted only univariable regressions for the stratified analysis, as the number of ZCTAs in each stratified model was low and multivariable analyses would be overfit.

## Results

Descriptive summaries of the covariates of interest and vaccine accessibility and coverage are included in Table 1. Figure 1 shows the location of vaccination sites across King County for both school age and adolescent groups. There were nearly three times as many adolescent versus school age vaccination sites (434 vs 152). For both groups, most sites were clustered around Seattle (on the western side of the county) and the immediate neighboring suburbs. The eastern region of King County, WA is more mountainous and contains less dense, more spread-out communities. Notably, there were almost no school age vaccination sites listed in the Vaccine Locator for eastern King County, while there were a small number of adolescent sites. Vaccine accessibility is shown in figure 2. The interquartile range was 112.5 to 141.7 sites per 100,000 individuals for adolescents and 31.6 to 42.9 sites per 100,000 individuals for school age children. The highest areas of accessibility were in southern Seattle and the nearby suburbs of Kent, Renton, and Mercer Island. Figure 3 shows vaccine coverage in the region. The interquartile range for adolescents was 64.3% to 95% (the maximum reported value by the PHSKC website) and for school age children it was 35.2% to 69.3%. Again, coverage was highest in Seattle and its neighbors. However, compared to accessibility, the ZCTAs with the highest coverage were in northern Seattle and the suburbs of Bellevue, Kirkland, and Redmond.

**Figure 1:**
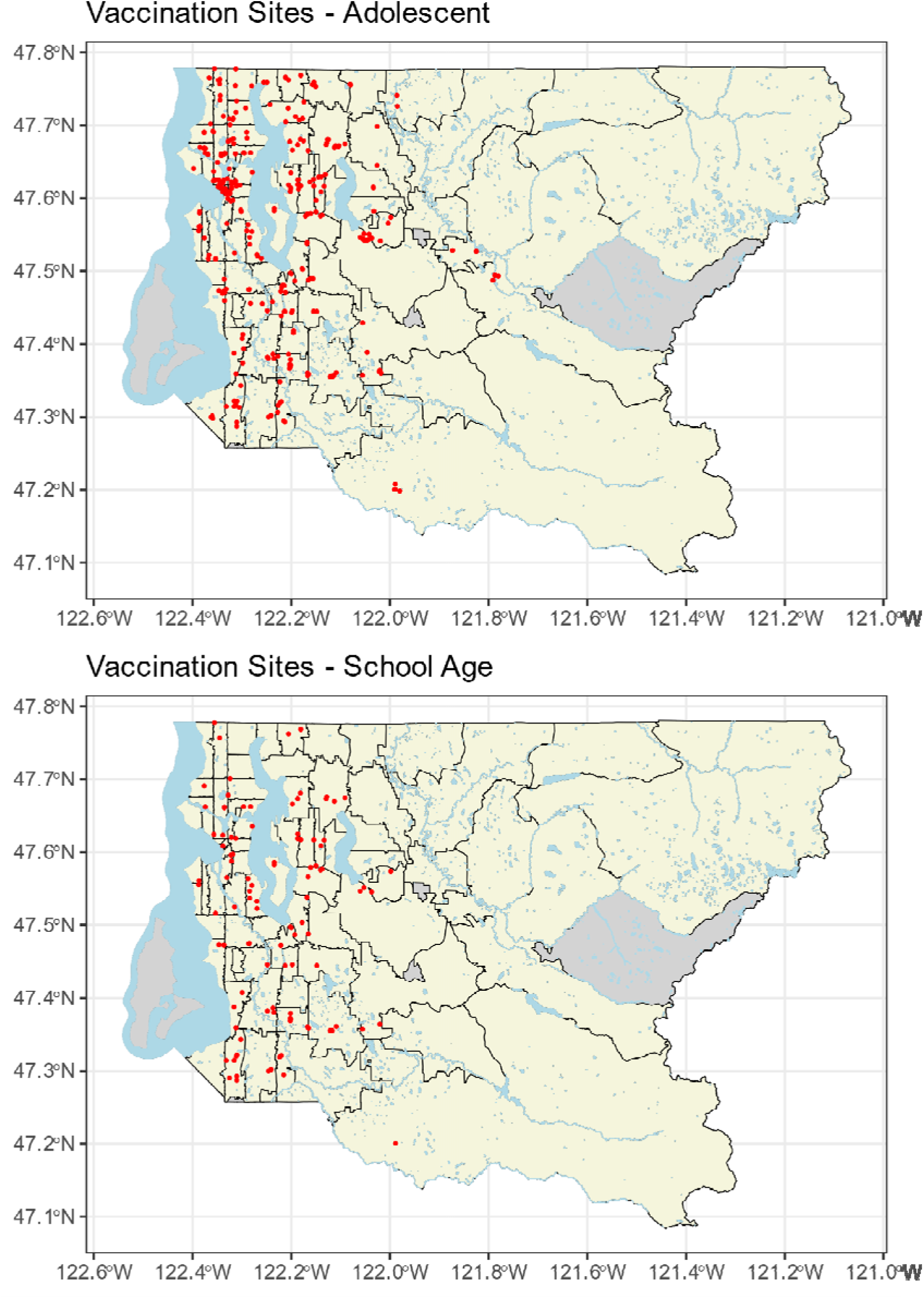
Vaccination sites within King County separated by age. Each red dot represents one site. Black lines indicate ZCTA boundaries. Gray ZCTAs were not included in the analysis.

**Figure 2:**
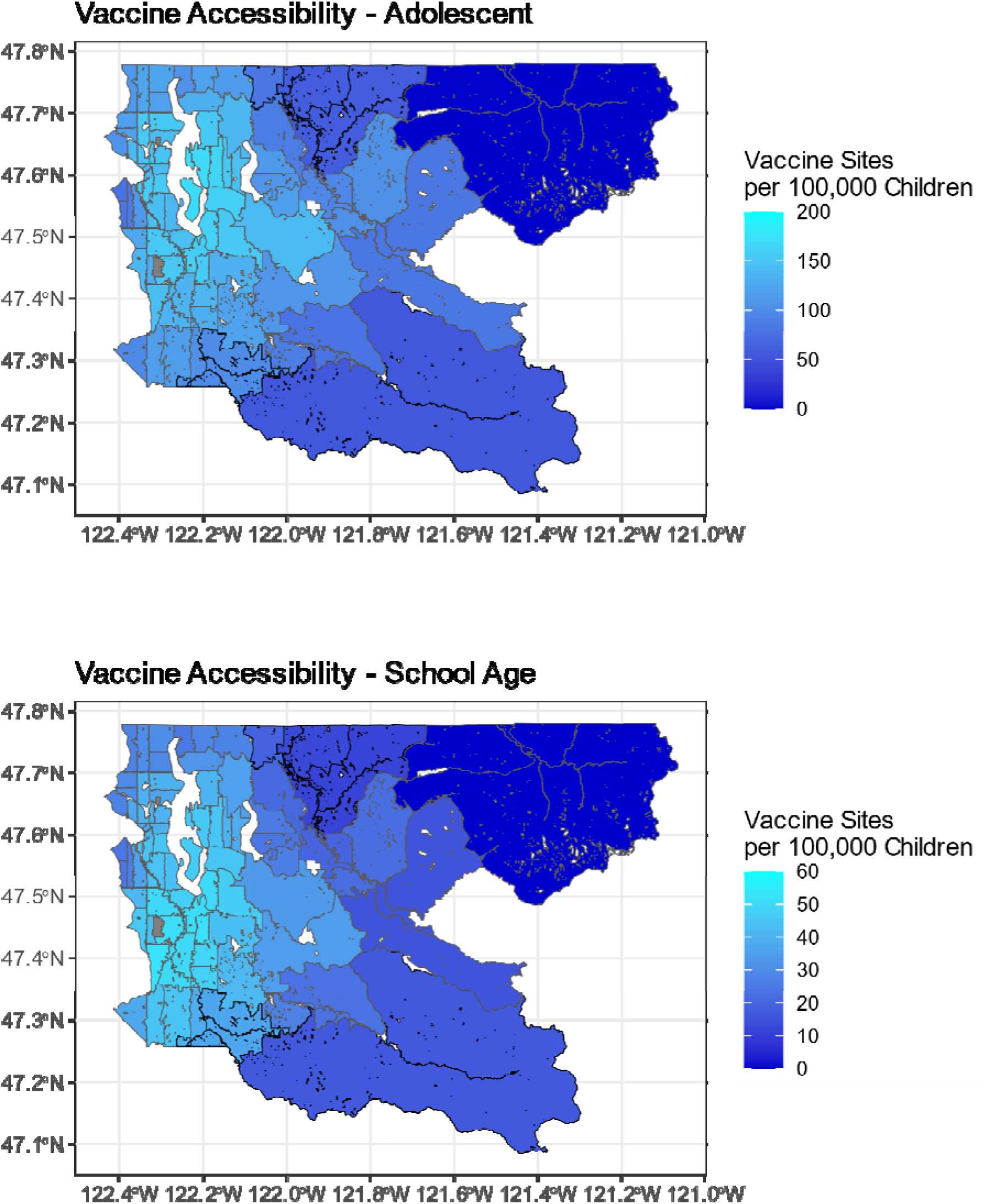
Vaccine accessibility by ZCTA in King County. Note significantly different scale magnitudes (maximum score of 200 for adolescents vs 60 for school aged children) in order to better compare the trends across maps. Gray ZCTAs were excluded due to zero population (University of Washington Main Campus Buildings and Seattle-Tacoma International Airport)

**Figure 3:**
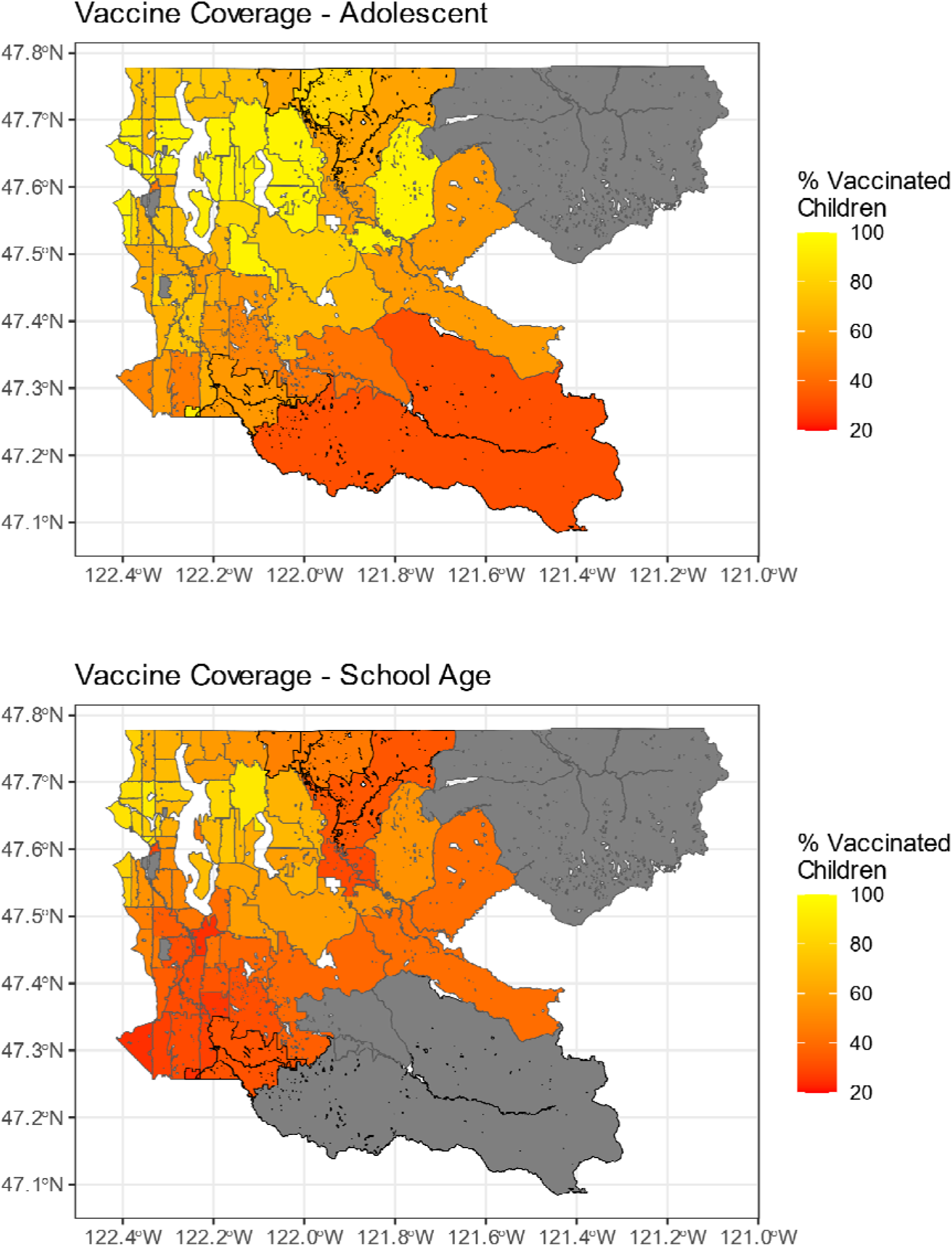
Vaccine coverage by ZCTA in King County. Gray ZCTAs were censored due to less than 10 residents of the age group.

**Table 1:**
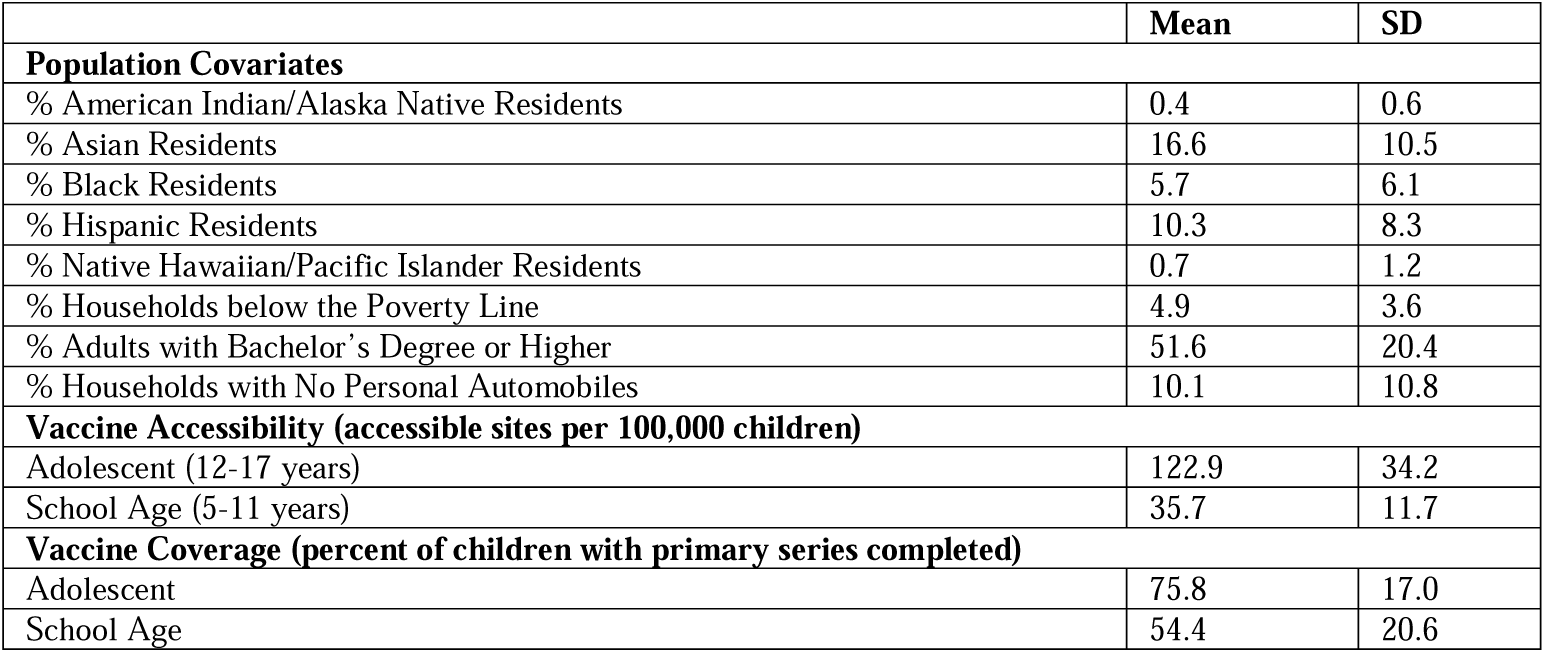
Descriptive statistics of King County ZCTAs.

There was a significant difference in vaccine accessibility between adolescents and school aged children across ZCTAs. There was a mean difference of 87.8 fewer vaccine sites per 100,000 children for school age children compared to adolescents (p< 0.001). Similarly, there was a significant mean difference in vaccine coverage with school age children having 21.5% lower coverage than adolescents (p< 0.001). Despite the differences in magnitude of vaccine accessibility and coverage between the two age groups, they were very strongly correlated to each other. Accessibility was correlated across ages with an R of 0.901 and coverage was correlated with an R of 0.786. Regression analyses to determine what covariates are associated with vaccine accessibility are shown in Table 2. The relationships identified are the same across school age and adolescent vaccine accessibility. The percent of adults with a bachelor’s or higher degree within a ZCTA and the percent of Asian residents were positively associated with the ZCTA’s accessibility.

**Table 2:**
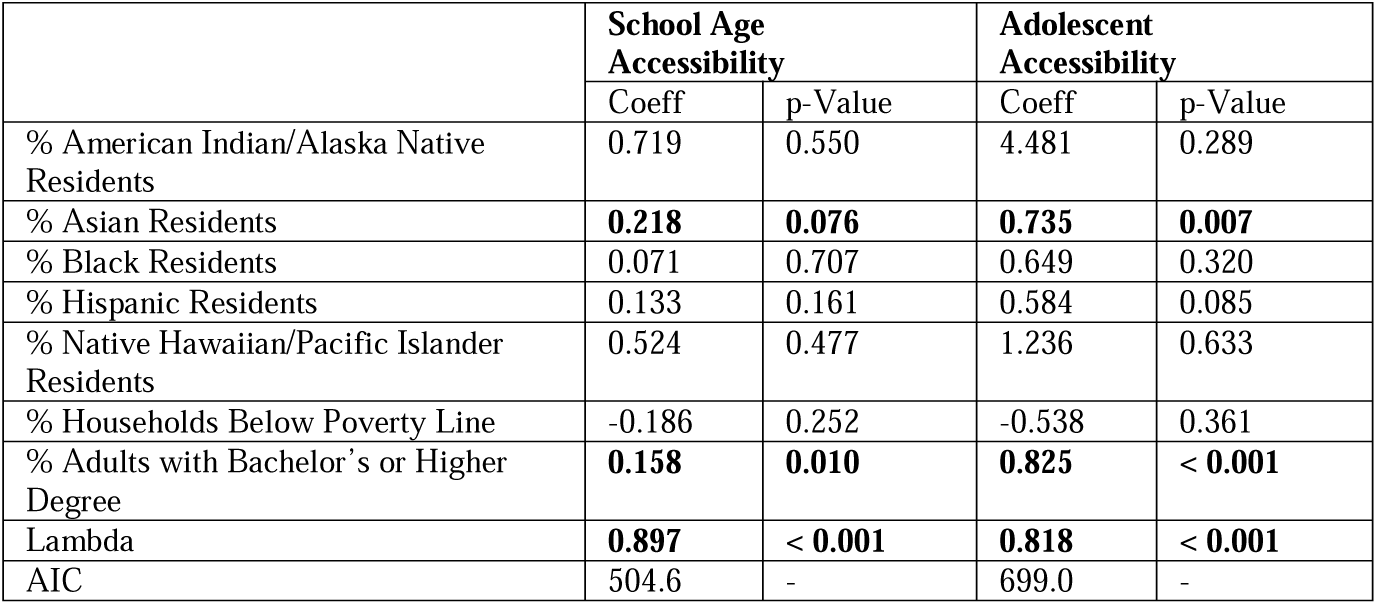
Regression models of vaccine accessibility. Lambda is spatial autoregressive coefficient in spatial error term. Terms with significance at p < 0.05 are bolded.

Regression analyses to evaluate associations with vaccine coverage are shown in Table 3. For both school age children and adolescents, vaccine accessibility was significantly associated with a higher percent of vaccinated children. However, this relationship was non-significant for school age children in the multivariate analyses. For adolescents, the multivariate analyses showed the effect of vaccine accessibility interacted significantly with educational attainment, with a positive relationship for both accessibility and educational attainment and negative effect for the interaction of the two variables. Therefore, adolescent vaccine coverage is predicted to increase with accessibility or educational attainment increase while the other remains low. However, when educational attainment is already high, accessibility was no longer predicted to cause a significant increase in coverage. Based on these results, we re-stratified the analysis on percent of adults with bachelor’s degrees or higher, as shown in table 4. Vaccine accessibility was only significantly associated with coverage among adolescents living in ZCTAs with < 50% bachelor’s degrees among adults and no relationship between accessibility and coverage in the ≥ 50% bachelor’s degrees ZCTAs.

**Table 3:** Regression models of vaccine coverage. Lambda is spatial autoregressive coefficient in spatial error term. Terms with significance at p < 0.05 are bolded. ^†^Univariate Lambda and AIC is for the Vaccine Access models.

| Adolescent<br>Vaccination Coverage | Univariate |  | Multivariate |  | Multivariate with<br>Interaction |  |
| --- | --- | --- | --- | --- | --- | --- |
|  | Coeff | p-Value | Coeff | p-Value | Coeff | p-Value |
| Vaccine Access | **0.231** | **0.005** | -0.061 | 0.261 | **0.385** | **0.013** |
| % American Indian/Alaska Native Residents | **-7.365** | **0.036** | -2.523 | 0.251 | -0.715 | 0.735 |
| % Asian Residents | **0.592** | **0.003** | **0.351** | **0.011** | **0.283** | **0.026** |
| % Black Residents | -0.302 | 0.342 | 0.394 | 0.176 | 0.058 | 0.842 |
| % Hispanic Residents | -0.754 | 0.057 | **1.042** | **0.003** | **0.823** | **0.016** |
| % Native Hawaiian/Pacific Islander Residents | 2.073 | 0.296 | **4.175** | **< 0.001** | **4.777** | **< 0.001** |
| % Households Below Poverty Line | **-1.228** | **0.032** | -0.816 | 0.118 | -0.656 | 0.186 |
| % Adults with Bachelor's Degree or Higher | **0.629** | **< 0.001** | **0.935** | **< 0.001** | **1.896** | **< 0.001** |
| Access-Bachelor's Degree Interaction Term | - | - | - | - | **-0.007** | **0.002** |
| Lambda | **0.556<sup>†</sup>** | **< 0.001** | -0.134 | 0.466 | -0.217 | 0.240 |
| AIC | 620.9<sup>†</sup> | - | 578.97 | - | 572.4 | - |
| School Age<br>Vaccination Coverage | Univariate |  | Multivariate |  | Multivariate with<br>Interaction |  |
|  | Coeff | p-Value | Coeff | p-Value | Coeff | p-Value |
| Vaccine Access | **0.686** | **0.006** | 0.074 | 0.670 | 0.505 | 0.234 |
| % American Indian/Alaska Native Residents | **-6.541** | **0.029** | **-4.106** | **0.039** | -3.546 | 0.079 |
| % Asian Residents | **0.485** | **0.006** | **0.412** | **0.002** | **0.370** | **0.006** |
| % Black Residents | -0.460 | 0.278 | 0.222 | 0.457 | 0.215 | 0.467 |
| % Hispanic Residents | **-0.719** | **0.039** | **0.652** | **0.013** | **0.611** | **0.022** |
| % Native Hawaiian/Pacific Islander Residents | -2.571 | 0.142 | -0.733 | 0.537 | -0.873 | 0.463 |
| % Households Below Poverty Line | **-1.601** | **< 0.001** | **-1.145** | **0.002** | **-1.158** | **0.001** |
| % Adults with Bachelor's Degree or Higher | **0.909** | **< 0.001** | **0.811** | **< 0.001** | **1.127** | **< 0.001** |
| Access-Bachelor's Degree Interaction Term | - | - | - | - | -0.009 | 0.260 |
| Lambda | **0.845<sup>†</sup>** | **< 0.001** | **0.781** | **< 0.001** | **0.752** | **< 0.001** |
| AIC | 600.0<sup>†</sup> | - | 546.0 | - | 546.8 | - |

**Table 4:**
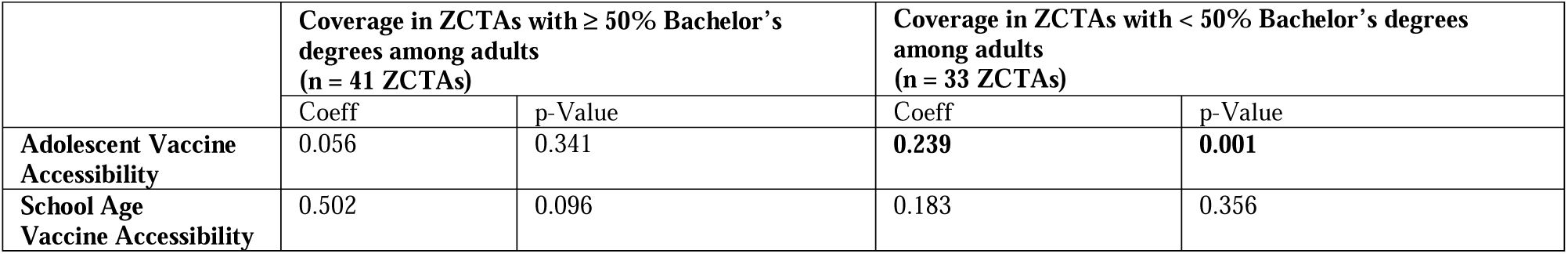
Stratified spatial regression analysis of the relationship between vaccine accessibility and coverage, stratifying on age group and the educational attainment of the ZCTAs. No other covariates were included in these analyses. Terms with significance at p < 0.05 are bolded.

| | Coverage in ZCTAs with $\geq 50\%$ Bachelor's degrees among adults<br>(n = 41 ZCTAs) | | Coverage in ZCTAs with $< 50\%$ Bachelor's degrees among adults<br>(n = 33 ZCTAs) | |
| --- | --- | --- | --- | --- |
|  | Coeff | p-Value | Coeff | p-Value |
| Adolescent Vaccine Accessibility | 0.056 | 0.341 | **0.239** | **0.001** |
| School Age Vaccine Accessibility | 0.502 | 0.096 | 0.183 | 0.356 |

For the other covariates, percentage of adults with a bachelor’s or higher degree, percent of Asian residents, and percent of Hispanic residents were found to be significantly associated with increased vaccine coverage in both age groups. Additionally, in the adolescent analysis, the percent of Native Hawaiian/Pacific Islander (NH/PI) was positively associated with coverage. However, both the Hispanic and NH/PI percentage effects had inconsistent effects when compared with the univariable models. NH/PI percentage did not have a significant effect, and Hispanic percentage had a statistically significant negative association. The percent of households whose income was below the poverty line and the percentage of American Indian/Alaska Native (AI/AN) residents were significantly negatively associated with coverage in both age groups in the univariable models, but they were only significantly associated in the school age group in the multivariable model.

## Discussion

In this study we used the E2SFCA method to estimate pediatric COVID-19 vaccine accessibility in King County, WA and investigated differences in access and coverage across different age groups and other socioeconomic and demographic variables. At the time of writing of this article, there have been several papers recently published mapping COVID-19 vaccine accessibility, but none that have focused specifically on pediatric populations.^18–20^ We showed that the areas of highest accessibility within King County were located in Seattle and the immediately adjacent suburbs. In particular, the southernmost neighborhoods of Seattle and the suburbs of Renton, Newcastle, and Mercer Island had the highest accessibility scores. This is likely due to the confluence of multiple interstate highways (I-5, I-90, and I-405) in this area which decrease the estimated travel times to vaccination sites.

While the relative patterns of vaccine accessibility are similar between the two age groups, the magnitude was found to be significantly different, with most tracts having school age accessibility scores that were 1/3^rd^ to 1/4^th^ of the adolescent score. This reflects the lower number of vaccination sites providing school age vaccines. Similarly, vaccine coverage was shown to have the same relative patterns between the two age groups, but there was a mean difference of −21.5% between adolescents and school age children coverage rates. Unfortunately, this is in line with the US national trends on pediatric vaccination. By the end of August 2022, 60.4% of US adolescents had completed the primary COVID-19 vaccination series while only 30.5% of school age children had.^21^ This likely in part reflects the differences in eligibility and authorization timeline between the two age groups, with the vaccine for 12 to 15-year-olds authorized on 5/10/2021 in the United States, while the vaccine for 5 to 11-year-olds were authorized on 10/29/2021. Furthermore, the dosage differences for the school age group from the adolescent/adult vaccine may have required vaccination sites to develop different vaccine storage protocols and have additional staffing requirements to meet the additional demand.

Given that pediatric COVID-19 vaccination lagged compared to adult coverage, it was important to identify significant predictors of vaccination coverage. The most notable difference between the school age and adolescent groups is that vaccine access was not associated with coverage in school age children in any of the multivariate analyses, while it was associated for adolescents after adjusting for an interaction term with our measure for neighborhood educational attainment (percentage of adults with bachelor’s or higher degrees). Interestingly, the relationship between accessibility and coverage appears to be limited to ZCTA with lower levels of educational attainment based on the stratified analysis in Table 4. In both age groups, average educational attainment was the single strongest positive predictor of both coverage rates and accessibility scores at the ZCTA level. There is evidence showing that higher levels of educational attainment have been associated with higher interest in COVID-19 vaccination in several survey studies in the United States and several other countries.^22–25^ One hypothesis to explain why accessibility is a less important predictor in neighborhoods with higher levels of educational attainment may be that caretakers who have higher levels of educational attainment may have a greater surplus of time and more flexible work schedules to take their children to be vaccinated. Therefore, their likelihood to be vaccinated could be less dependent on the spatial accessibility of vaccination sites. Alternatively, accessibility was correlated with educational attainment for both age groups. It may be that there is a specific threshold of spatial accessibility that is necessary for high rates of vaccination coverage, and the likelihood that a ZCTA has sufficient accessibility is better predicted by educational attainment. Accessibility beyond that level has diminishing returns, and therefore results in a statistically non-significant relationship in our models.

Racial disparities in COVID-19 outcomes and vaccination have been a major health equity concern throughout this pandemic. One previous study looking at COVID-19 vaccination coverage in Texas found that ZCTAs with higher than the median percentage of Black, Hispanic, and American Indian/Alaska Native residents had significantly lower rates of vaccine coverage.^26^ Our analysis found that the percentage of AI/AN residents was associated with lower rates of school age vaccine coverage in the multivariate model without interactions, but this relationship was non-significant in the model with interactions. Meanwhile, the percent Asian, Hispanic, and NH/PI residents were all associated with higher levels of vaccine coverage. It is important to highlight that this is an ecological study, and therefore the analysis occurs at the level of ZCTAs rather than individuals. Therefore, the relationships found for racial composition should not be interpreted as equivalent relationships for individuals of that particular race, as this interpretation would represent an ecological fallacy. Instead, the goal of including racial composition variables was to look for possible effects of structural racism and segregation. To illustrate this point, the ecological relationships between race/ethnicity composition of a ZCTA and vaccine coverage shown here differ from published individual-level data on race and vaccination by PHSKC on their COVID-19 vaccination dashboard.^12^ Vaccine coverage among AI/AN individuals across all ages (including adults) in King County was > 95% in July 2022, while Hispanic and Black individuals had lower rates of coverage at 70.1% and 76.7% respectively. This difference with our ZCTA level results highlights the difference between ecological and individual relationships. It is also important to call out the potential effect of measurements with small numbers with limited variability. Within ZCTAs, AI/AN and NH/PI percentage was low across the region with the mean percentage less than 1%, so these variables’ relationships are at higher risk for bias from influential observations. For Hispanic percentage, the true relationship is difficult to determine given the inconsistent estimated effects between the multivariable and univariable models. Percentage of household poverty was also negatively associated with school age vaccination rates which is consistent with previous work looking at vaccination coverage and healthcare utilization during the pandemic.^8,26^

This study does have several important limitations. This is a cross-sectional analysis which limits us from establishing any causal inferences. As with any spatial analyses, the modifiable areal unit problem may affect our ability to properly identify relationships.^27^ Spatial relationships can be biased by how administrative units such as ZCTAs are drawn, because the boundaries affect the aggregation of data. ZCTAs were the only sub-county level of data we were able to access for vaccine coverage. We also limited our study to vaccination sites within King County due to our access of vaccine coverage data, and we did not account for intercounty travel in calculating accessibility. Another limitation is that our measures of vaccine accessibility are estimates based on assumptions of travel and healthcare utilization behaviors. We have formulated them using existing street network data, publicly available vaccine site data, and existing literature and data on healthcare travel behaviors. The severe disruptions of the COVID-19 pandemic to day-to-day travel and the staggered nature of vaccine rollouts make it difficult to predict the lived experience of the individuals living in these neighborhoods when accessing the vaccine. Finally, we were not able to account for the effects of the mobile and school vaccination sites, tribal sites, and community vaccine drives that were utilized by King County to help provide the vaccine.^28^ These vaccine drives outside of traditional clinics and pharmacies were an important tool used by PHSKC, other health agencies, and community leaders to help expand coverage to specific communities, especially for communities that have been historically underserved.^29,30^

Despite these limitations, we believe this analysis can be useful in evaluating pediatric COVID-19 vaccination access to help guide public health activities to improve rates of the pediatric population who are up to date with recommended COVID-19 vaccinations. Vaccine access and coverage for the school age population was significantly lower than for adolescents. More vaccination sites are needed to provide doses across the full age range of pediatric patients. Furthermore, there should be targeted efforts in neighborhoods with lower levels of educational attainment and higher rates of household poverty to increase vaccine coverage in these ZIP codes. These also may be areas that would benefit the most from future mobile vaccination sites as they had the lowest accessibility scores as well. This study evaluated several publicly available community-level predictors of vaccine access and coverage. However, this cannot substitute for a nuanced community-driven understanding of vaccine perception and barriers to access. It is important for researchers, healthcare providers, public health organizations, and health systems to partner with local communities to ensure high and equitable coverage of vaccines. Ultimately, increasing COVID-19 vaccination rates and re-vaccinating with updated formulations among children remains an important priority for pediatric healthcare providers and public health workers to help prevent future waves and outbreaks.

## Supporting information

Supplement 1: E2SFCA Technical Details

## Data Availability

Current vaccine locations can be found using the Vaccinate WA website at: https://vaccinelocator.doh.wa.gov/?language=en and updated vaccine coverage data can be found using the Public Health Seattle & King County website at: https://kingcounty.gov/en/legacy/depts/health/covid-19/data/vaccination.aspx. These websites do not archive previous data. The specific datasets used in the analysis can be obtained at request of the authors and will be uploaded to a code repository once the paper is fully published.

## Acknowledgments

We would like to thank Daniel Casey and Sargis Pogosjan from PHSKC for discussions providing background regarding King County’s COVID-19 vaccination effort and for reviewing this manuscript prior to submission.

## Role of the funding source

There was no specific funding source involved in the design, analysis, or writing of this paper.

## Declarations of Interest

We declare no competing interests.

## Data Sharing

Current vaccine locations can be found using the Vaccinate WA website at: https://vaccinelocator.doh.wa.gov/?language=en and updated vaccine coverage data can be found using the Public Health Seattle & King County website at: https://kingcounty.gov/en/legacy/depts/health/covid-19/data/vaccination.aspx. These websites do not archive previous data. The specific datasets used in the analysis can be found in our code repository at [repository link to be added at time of resubmission]

## Notes

### Competing Interest Statement

The authors have declared no competing interest.

