## Supplement 1: E2SFCA Technical Details for "Investigating the spatial accessibility and coverage of the pediatric COVID-19 vaccine: an ecologic study of regional health data"

**Supplement 1: Calculation of time weights and implementation of E2SFCA**

The Enhanced Two-Step Floating Catchment Area (E2SFCA) is an extension of the traditional Two-Step Floating Catchment Area (2SFCA), which involves two primary steps.^1^ The first step of 2SFCA is to calculate the population ratio ($R_{j}$) of each vaccination site $j$ by adding up the population ($P_{k}$) of all ZCTAs $k$ that fall within the catchment area based on the travel time between the population center and site ($t_{kj}$):

$$R_{j}=\frac{1}{\sum_{k\in\{t_{kj}<t_{0}\}} P_{k}}$$

where $t_{0}$ represents the maximum catchment distance in travel time. In the original formula for this method, the numerator can be represented as $S_{j}$ which represents the quantity of service provided at a site (e.g. number of vaccines available, number of providers giving vaccines). However, the original dataset did not provide the number of vaccines or providers attached to a site, so we simplified it to the case where $S_{j}$ = 1.

The second step is to calculate each ZCTA’s ($i$) accessibility score $A_{i}$ by summing all the ratios that fall within the catchment range of an individual seeking the vaccine ($t_{ij}<t_{0}$):

$$A_{i}=\sum_{i\in t_{ij}<t_{0}} R_{j}=\sum_{i\in t_{ij}<t_{0}} \frac{1}{\sum_{k\in\{t_{kj}<t_{0}\}} P_{k}}$$

For our scenario, $A_{i}$ can be thought as the number of vaccination sites per capita in that ZCTA. For easier legibility on the maps and tables, we multiply it by 100,000 so the final units are “vaccination sites per 100,000 children.”

The E2SFCA then enhances 2SFCA by adding weights based on the distance for both steps. In Luo and Qi’s original E2SFCA paper, they enhance 2SFCA by dividing the floating catchment areas into three different areas and assigning each a weight score using a gaussian distribution.^1^ The catchment with the smallest distance would have the highest weight and the catchment with the largest distance would have the lowest.

We take a different approach in our implementation. Rather than assign *a priori* discrete catchment ranges, we used existing data from the 2017 National Household Travel Survey to make an empiric cumulative density function (eCDF) of travel time for healthcare related trips. Broader analysis of this dataset was a subject of a separate paper,^2^ but we will focus on discussion of the eCDFs here. The eCDF is calculated as a simple step function with no smoothening. We separated the eCDFs based on automobile use ($E_{C}(t)$) and public transit use ($E_{T}(t)$) given significant differences in the distribution. We then defined the weight function of each transportation mode as: $W\left( t \right)=1-E\left( t \right)$.

There are two major benefits in this approach. First, when the original E2SFCA paper was developed, calculating the travel time between every possible combination of provider and population site was extremely time intensive and impractical. As a result, discrete catchment ranges were used. However, with the advancements in computing power, we can now calculate every pair in a reasonable time frame. For reference, our code can be run on a high-end consumer-grade desktop computer (R 4.2.1, AMD Ryzen 3700X, 64 GB of DDR4 RAM, Windows 11) in 10 to 12 hours on average. Since these travel times are continuous, we can calculate more precise estimates of accessibility. The second benefit of this approach is that instead of assuming a Gaussian distribution for the weighting we can use real-world data specifically looking at healthcare related travel. The functions can be seen below:

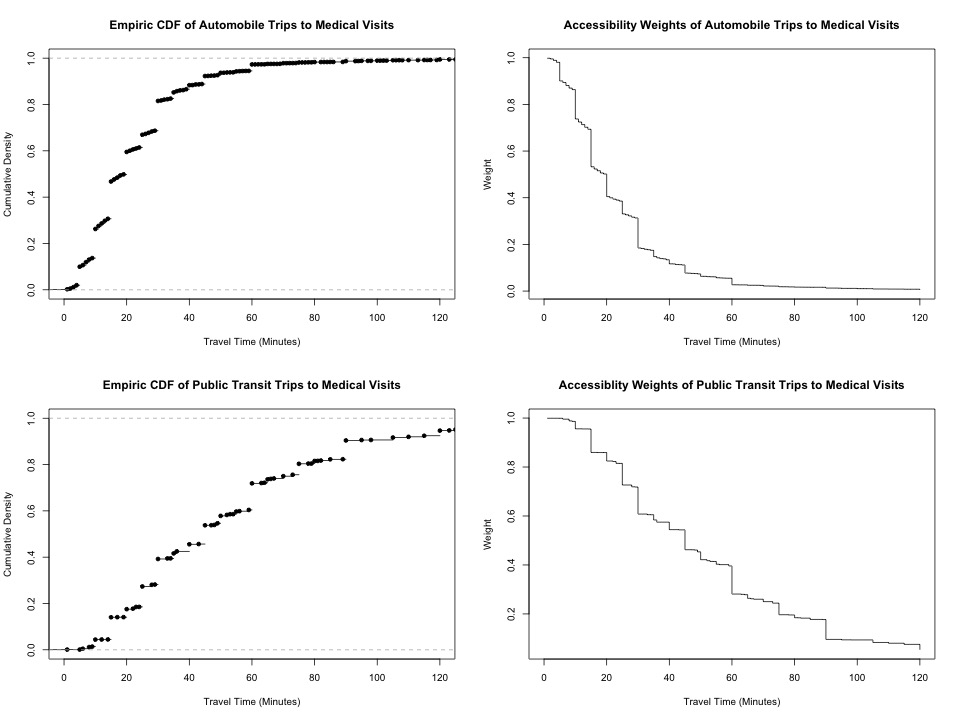

We then integrate these weight functions into the original 2SFCA equations in a similar manner to Luo and Qi. For the population ratio step, we only incorporate automobile travel times $(t_{kj}^{C})$ and weights $(W_{C}(t))$ in order to maximize the amount of population within the catchment area that each site covers. The revised equation is as follows:

$$R_{j}=\frac{1}{\sum_{k} W_{C}(t_{kj}^{C})P_{k}}$$

For the second step we stratify the accessibility score calculation by transportation mode (public transit, $A_{i}^{T}$, versus automobile, $A_{t}^{C}$). We then calculate a weighted average of the two accessibility scores based on the percent of households with personal automobile access in each ZCTA ($c_{i}$). The full equation is below:

$$A_{i}=c_{i}\times A_{t}^{C}+\left( 1-c_{i} \right)\times A_{t}^{T}$$

$$A_{i}=c_{i}\sum_{j} W_{c}(t_{ij}^{C})\times R_{j}+(1-c_{i})\sum_{j} W_{T}(t_{ij}^{T})\times R_{j}$$

$$A_{i}=c_{i}\sum_{j} W_{c}(t_{ij}^{C})\times\frac{1}{\sum_{k} W_{C}(t_{kj}^{C})P_{k}}+(1-c_{i})\sum_{j} W_{T}(t_{ij}^{T})\times\frac{1}{\sum_{k} W_{C}(t_{kj}^{C})P_{k}}$$

where $W_{T}$ is the weight function for public transit and $t_{ij}^{T}$is the public transit travel time. It is important to note that these calculations are also stratified by age group (school age vs adolescent), and that $P_{k}$is the population size for the specific age group.
